# Breakthrough Symptomatic COVID-19 Infections Leading to Long Covid: Report from Long Covid Facebook Group Poll

**DOI:** 10.1101/2021.07.23.21261030

**Authors:** Daisy Massey, Diana Berrent, Harlan Krumholz

## Abstract

Vaccines have been shown to be extremely effective in preventing COVID-19 hospitalizations and deaths. However, a question remains whether vaccine breakthrough cases can still lead to Post-Acute Sequelae of SARS-CoV-2 (PASC), also known as Long Covid. To address this question, the Survivor Corps group, a grassroots COVID-19 organization focused on patient support and research, posted a poll to its 169,900 members that asked about breakthrough cases, Long Covid, and hospitalizations. 1,949 people who self-report being fully vaccinated have responded to date. While robust data are needed in a larger, unbiased sample to extrapolate rates to the population, we analyzed the results of this public poll to determine what people were reporting regarding Long Covid after breakthrough infection and to prompt discussion of how breakthrough cases are measured. The poll was posted in the Survivor Corps Facebook group (∼169,900 members). Of the 1,949 participants who responded to the poll, 44 reported a symptomatic breakthrough case and 24 of those reported that the case led to symptoms of Long Covid. 1 of these 24 cases was reported to have led to hospitalization in addition to Long Covid.

## Introduction

While the current COVID-19 vaccines are highly effective at preventing hospitalizations and deaths, some breakthrough infections of SARS-CoV-2 have been found to occur.^1^ As of April 30, 2021, 10,262 SARS-CoV-2 vaccine breakthrough infections had been reported to the Center for Disease Control.^2^ A remaining question is to what degree breakthrough infections are associated with debilitating and long-lasting symptoms, or Long Covid. In the unvaccinated population, both hospitalized and non-hospitalized people with SARS-CoV-2 infections have been reported to develop Long Covid.^3^ Although the rate of Long Covid among people with COVID-19 who are not hospitalized is still unknown, current evidence indicates that it may be between 14% and 30%.^3,4^

To produce information related to reinfection and Long Covid, Survivor Corps, a COVID-19 grassroots organization dedicated to patient support and research, posted a poll in its public Facebook group of ∼169,900 members.^5,6^ The poll, posted on June 2, 2021, asked participants to share their COVID-19 infection status post-vaccine, including whether participants had developed Long Covid or been hospitalized.

Accordingly, using the public information that Survivor Corps collected we sought to organize it and preprint it to give voice to patient reports, give the public easy access to it, and enable it to be archived and citable. Our specific aim was to show what the highly selected group of respondents reported regarding the development of Long Covid from symptomatic infection after being fully vaccinated, which we term breakthrough infection. Our goal is to stimulate discussion regarding this topic and to stimulate future research efforts.

## Methods

Survivor Corps, an organization of people with Long Covid focused on patient support and research, aimed to produce information about breakthrough infections and Long Covid from their Facebook group. Thus, the group conducted a poll within its public Facebook group, which includes ∼169,900 people, about people’s experience after receiving the Pfizer-BioNTech, Moderna, or Johnson & Johnson/Janssen COVID-19 vaccine (Figure). The Facebook group is public and there is no requirement for signing up. Its stated purpose is to mobilize and connect COVID-19 survivors with the medical, scientific, and academic research community.

**Figure.**
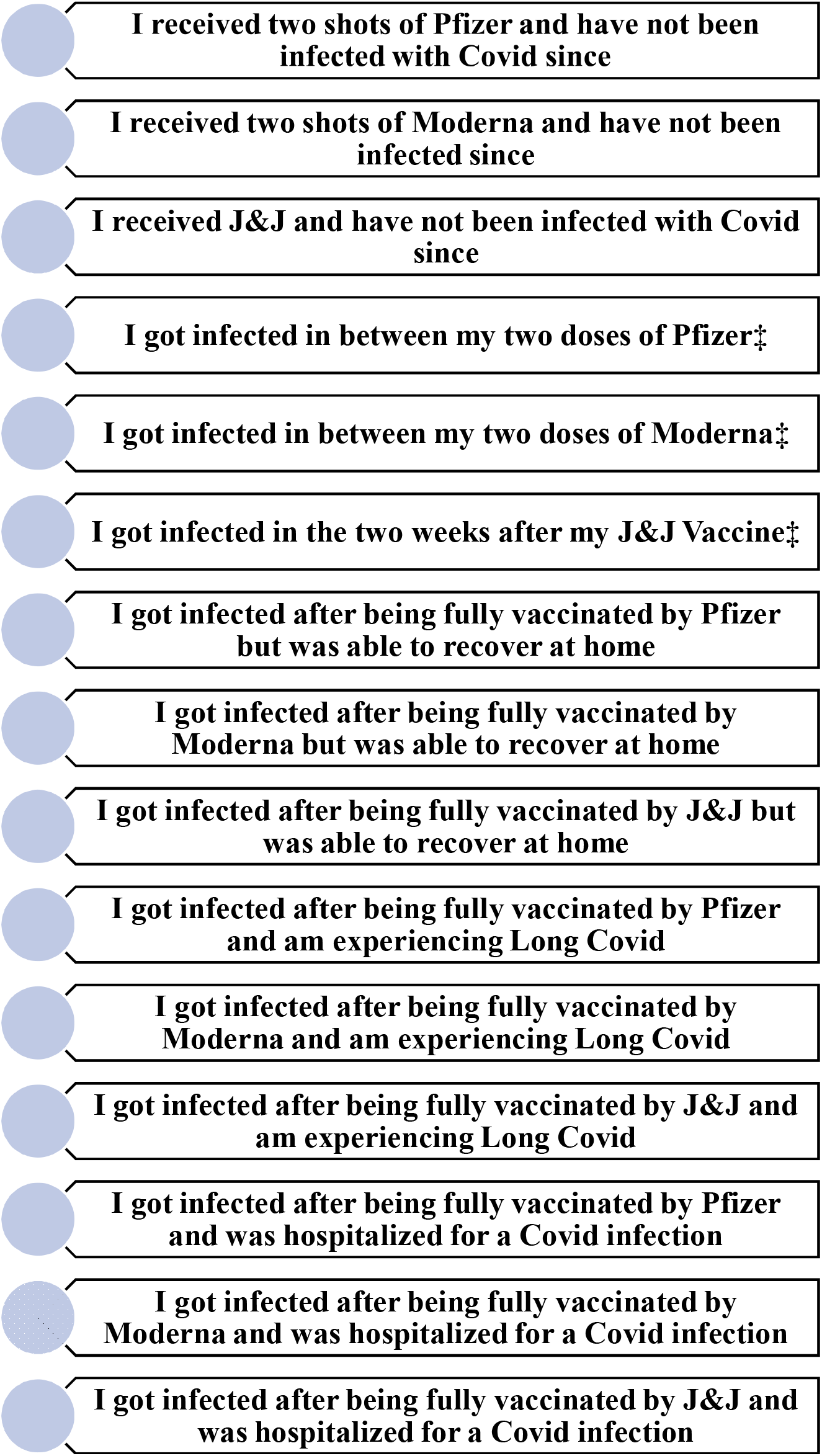
Survivor Corps Facebook group post-infection vaccine poll (Posted on June 2, 2021) ‡Indicates answer choices included in total of participants fully vaccinated, or fully vaccinated by vaccine, but not included in total of breakthrough cases.

The poll included options for whether people had been fully vaccinated by one of the vaccines and either remained uninfected, became infected with COVID-19 but recovered from symptoms, became infected with COVID-19 and developed Long Covid, or became infected with COVID-19 and were hospitalized (Figure). This poll focuses on responses that were relevant to breakthrough infections and Long Covid. Although the poll is still ongoing, the data below represent the findings from 1,949 members who reported being fully vaccinated and who participated in the poll by July 22, 2021.

Survivor Corps shared deidentified data with authors for the purpose of this report. Also, the data are publicly available in the Facebook group. The public nature of the data and the fact that the dataset that was shared was deidentified obviated the need for informed consent.

The number of breakthrough cases were reported overall and stratified by vaccine received. People who reported a breakthrough case and recovering fully at home were included in the breakthrough case total. People who reported becoming infected with COVID-19 before 2 weeks after the last vaccine dose were not included in the breakthrough case total as they were not fully vaccinated at the time of infection, but they were included in the sample. Those who reported an asymptomatic case, adverse reaction, or incomplete vaccination were excluded from the sample.

## Result

Of the 1,949 fully vaccinated poll participants, 44 reported a breakthrough case (Table 1). Of these, 24 reported that their breakthrough case led to Long Covid. 3 cases were reported to have led to hospitalization and 1 of these cases was also reported to have led to Long Covid. There were 1,024 poll participants who reported receiving the Pfizer-BioNTech vaccine, 775 the Moderna vaccine, and 150 the Johnson & Johnson/Janssen vaccine (Table 2). The number of breakthrough cases by vaccine was 19 among people who received Pfizer-BioNTech, 17 among people who received Moderna, and 8 among people who received Johnson & Johnson/Janssen.

**Table 1.** Rates of reported COVID-19 breakthrough infection among fully vaccinated people.

| Participants N=1949 |  | Number of Cases |
| --- | --- | --- |
| Symptomatic Breakthrough Cases N=44 |  | 44 |
|  | Breakthrough that led to hospitalization without Long Covid | 2 |
|  | All breakthrough that led to Long Covid | 24 |
|  | Breakthrough that led to Long Covid and hospitalization | 1* |

\*This case is included in the total number of breakthrough cases leading to Long Covid.

**Table 2.**
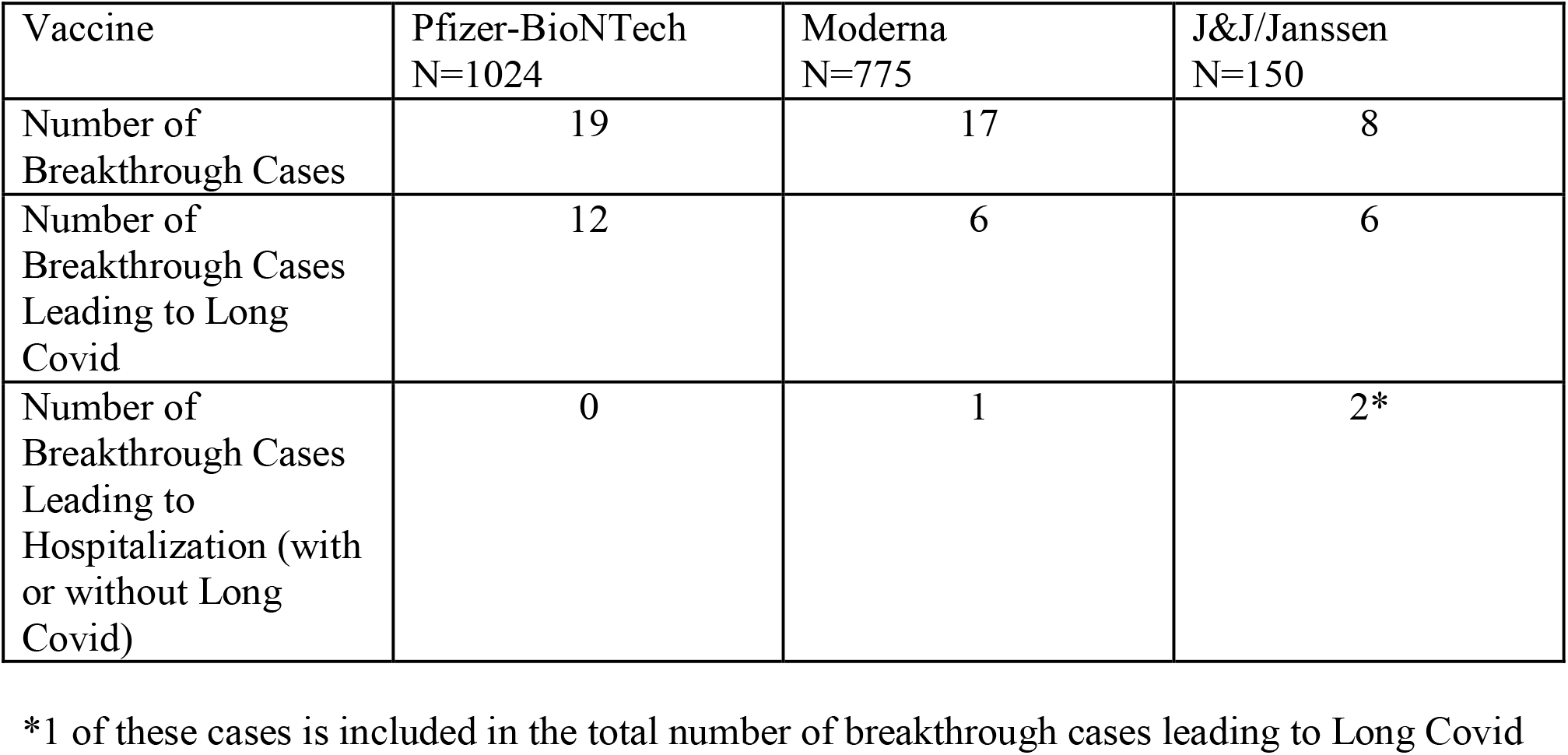
Rates of reported COVID-19 breakthrough infection among fully vaccinated by vaccine.

| Vaccine | Pfizer-BioNTech<br>N=1024 | Moderna<br>N=775 | J&J/Janssen<br>N=150 |
| --- | --- | --- | --- |
| Number of Breakthrough Cases | 19 | 17 | 8 |
| Number of Breakthrough Cases Leading to Long Covid | 12 | 6 | 6 |
| Number of Breakthrough Cases Leading to Hospitalization (with or without Long Covid) | 0 | 1 | 2* |

\*1 of these cases is included in the total number of breakthrough cases leading to Long Covid

## Discussion

The Survivor Corps poll found that some respondents with symptomatic breakthrough infections also reported Long Covid symptoms. Also, there was a respondent who reported that their breakthrough infection resulted in both Long Covid symptoms and a hospitalization. We report breakthrough by vaccine, but the nature of the poll precludes any conclusion about vaccine effectiveness.

The poll was limited and the respondents highly self-selected, so it is not possible to estimate rates of breakthrough or subsequent risk of Long Covid. The value of the survey is to highlight that some people report Long Covid following breakthrough and signals the importance of conducting more rigorous and detailed studies of whether risks of Long Covid after breakthrough are similar to what is being reported in initial infections. There is also a need for studies that identify risk factors for breakthrough infections and sequelae.

## Data Availability

All data are publicly available on the Survivor Corps Facebook page in the polls section.

https://www.facebook.com/groups/COVID19survivorcorps/posts/985579702190889

## Data Availability

All data are publicly available on the Survivor Corps Facebook page in the polls section.

https://www.facebook.com/groups/COVID19survivorcorps/posts/985579702190889

